# The incidence of COVID-19-related hospitalisation in migrants in the UK: Findings from the Virus Watch prospective community cohort study

**DOI:** 10.1101/2023.01.06.22283653

**Authors:** Wing Lam Erica Fong, Vincent G Nguyen, Rachel Burns, Yamina Boukari, Sarah Beale, Isobel Braithwaite, Thomas E Byrne, Cyril Geismar, Ellen Fragaszy, Susan Hoskins, Jana Kovar, Annalan MD Navaratnam, Youssof Oskrochi, Parth Patel, Sam Tweed, Alexei Yavlinsky, Andrew C Hayward, Robert W Aldridge

## Abstract

**Background:** Migrants in the UK may be at higher risk of SARS-CoV-2 exposure; however, little is known about their risk of COVID-19-related hospitalisation during waves 1-3 of the pandemic.

**Methods:** We analysed secondary care data linked to Virus Watch study data for adults and estimated COVID-19-related hospitalisation incidence rates by migration status. To estimate the total effect of migration status on COVID-19 hospitalisation rates, we ran fixed-effect Poisson regression for wave 1 (01/03/2020-31/08/2020; wildtype), and fixed-effect negative binomial regressions for waves 2 (01/09/2020-31/05/2021; Alpha) and 3 (01/06/2020-31/11/2021; Delta). Results of all models were then meta-analysed.

**Results:** Of 30,276 adults in the analyses, 26,492 (87.5%) were UK-born and 3,784 (12.5%) were migrants. COVID-19-related hospitalisation incidence rates for UK-born and migrant individuals across waves 1-3 were 2.7 [95% CI 2.2-3.2], and 4.6 [3.1-6.7] per 1,000 person-years, respectively. Pooled incidence rate ratios across waves suggested increased rate of COVID-19-related hospitalisation in migrants compared to UK-born individuals in unadjusted 1.68 [1.08-2.60] and adjusted analyses 1.35 [0.71-2.60].

**Conclusion:** Our findings suggest migration populations in the UK have excess risk of COVID-19-related hospitalisations and underscore the need for more equitable interventions particularly aimed at COVID-19 vaccination uptake among migrants.

## 1. Introduction

The coronavirus disease 2019 (COVID-19) pandemic highlighted and amplified existing health disparities among underserved populations in the UK. Migrants (non UK-born) often experience socio-economic and structural inequalities [1] that could contribute to an increased risk of severe acute respiratory syndrome coronavirus 2 (SARS-CoV-2) infection, and adverse COVID-19 outcomes [2], [3].

There are several unique risk factors that put migrants at higher risk of SARS-CoV-2 infection [4]. Migrants in the UK comprise a significant proportion of frontline workers, including healthcare, transport, and retail workers, who are at increased risk of SARS-CoV-2 infection and COVID-19 mortality [5]–[7]. Socio-economic disparities, such as living in overcrowded accommodation [8] and working in insecure and low-paid jobs [9], may impact the ability to take preventative measures including self-isolation, wearing personal protective equipment or social distancing [10]. Furthermore, healthcare access barriers [11] and limited linguistically and culturally appropriate public health messaging [12] may affect migrants’ awareness of the disease as well as of protective measures, putting them at higher exposure risk, and if infected, may result in later presentation at the hospital with more severe COVID-19 [13], [14].

Evidence across high-income countries (HICs) has demonstrated an increased risk of adverse COVID-19 outcomes among migrants compared to non-migrants. A systematic review of clinical outcomes for COVID-19 among migrant populations in HICs [15] reported greater COVID-19-related hospitalisation risk among migrants compared to native-born individuals in Italy, Denmark and Kuwait. In the UK, a greater all-cause excess mortality risk was observed in migrants compared to UK-born individuals during early 2020 [16]. Among UK healthcare workers who died of COVID-19 until late April 2020, migrants were also disproportionately represented (53%; 56/106) [17]. Some migrant groups have been found to be under-immunised for certain routine vaccines and recent studies show low intention to take up, and uptake of, the COVID-19 vaccine [3], [18]–[20], putting them at a higher risk of severe COVID-19 and hospitalisation. However, little is known about the effect of migration status on COVID-19-related hospitalisation during later pandemic phases in the UK.

To our knowledge, there are no large-scale studies in England that assessed the risk of COVID-19-related hospitalisation among migrants compared to UK-born individuals. Given the evidence for the increased risk of exposure to SARS-CoV-2 among migrants and their increased risk of COVID-19-related hospitalisation in other HICs countries, there is a need to estimate this risk in the UK across the different waves of the pandemic.

In this analysis we used data from the Virus Watch cohort study [21] linked to NHS hospital data to: 1) estimate the incidence of COVID-19-related hospitalisation across the first three COVID-19 infection waves in migrants in England compared to UK-born individuals and; 2) estimate the total (unadjusted and adjusted) effect of migration status on the rate of COVID-19-related hospitalisation among adults living in England during the three UK COVID-19 waves.

## 2. Methods

Virus Watch is a large prospective household cohort study of the transmission and burden of COVID-19 in England and Wales (n=58,497 as of March 2022). The study recruited participants from June 2020 to March 2022. Participants were self-selected into the study and only households with a lead householder able to speak English and access the internet were able to take part. The full study design and methodology has been described elsewhere [21], with relevant elements for the present study outlined here. The present study covered the following periods: from March 01 to August 31 2020 (wave 1 dominant variant: wildtype), from September 1 2020 to May 31 2021 (wave 2: Alpha), and from June 1 to November 30 2021 (wave 3: Delta).

### 2.1. Data sources and linkage

The primary source of data was the Virus Watch cohort linked to Hospital Episode Statistics (HES) Admitted Patient Care (APC), which contains details of all admissions at NHS hospitals in England and admissions to private or charitable hospitals paid for by the NHS [22]. Only completed consultations were used in the analysis (epistat=3). We also linked the Virus Watch dataset to the Second Generation Surveillance System (SGSS), the UK National Immunisation Management Service (NIMS) and mortality data from the Office for National Statistics (ONS). Linkage was conducted by NHS Digital with the linkage variables being sent in March 2021. Participant data were linked over time and between databases using the unique personal identifier recorded at all interactions with the NHS number, full name, date of birth and home address. The linkage period for HES encompassed data from March 2020 until November 2021, from March 2020 until August 2021 for SGSS Pillar 1 (testing NHS labs for patients in secondary care), from June 2020 until November 2021 for SGSS Pillar 2 (community testing), from October 2020 until December 2021 for NIMS, and from June 2020 until November 2021 for ONS mortality data.

### 2.2. Participants

Participants in this study were a subset of the Virus Watch study cohort. Participants were eligible for inclusion if they: 1) were ≥18 years upon cohort entry; 2) were registered with an English postcode; 3) were successfully linked to either HES, NIMS, SGSS or ONS mortality data, and; 4) reported a country of birth.

### 2.3. Exposure

The exposure was migration status (UK-born, Migrant) determined by self-reported country of birth at study enrolment. Participants who reported being born outside of the UK were identified as migrants. Comparisons were reported with the UK-born group as the reference category.

### 2.4. Outcome

The outcome was the number of COVID-19-related hospital admissions. We used ICD-10 codes U07.1 (confirmed COVID-19) and U07.2 (suspected/probable COVID-19) in HES APC data to identify COVID-19-related hospital admissions. Participants with less than one day difference between discharge date and next hospital admission date were counted to only have one hospital admission instead of two.

### 2.5. Covariates

Covariate adjustment for the primary analysis was informed by a directed acyclic graph (Supplementary Figure S1). The covariates considered were age group (18-44, 45-64, 65+ years), sex at birth, and binary minority ethnicity status (White British, ethnic minority). Missingness was present in the minority ethnicity status (0.3%, n=93) variable. Due to the small number of missing values, individuals with missing covariates were excluded from the analysis.

### 2.6. Other demographic and clinical characteristics

Participants were assigned a geographical region (ONS national regions) and a local area-level Index of Multiple Deprivation (IMD) quintile (1=most deprived, 5=least deprived) based on self-reported postcode of residence (MHCLG) [23]. Occupational risk was grouped using the Job Exposure Matrix for work-related COVID-19 exposure [24], classified as “Low”, “Elevated” and “High” occupational risk. Participants who reported to be unemployed or not working were categorised as “No occupational risk”. Clinical vulnerability was derived from self-reported data on immunosuppressive therapy, cancer diagnoses and chronic disease status. This was categorised as “Not clinically vulnerable”, “Clinically vulnerable” and “Clinically extremely vulnerable”. COVID-19 vaccination status (unvaccinated, 1 dose, 2 or more doses) was also determined by linking to NIMS data. Migrants’ self-reported date of entry into the UK was used to categorise into <1, 1-5 and 5+ years since entry in the UK upon follow-up for each wave. Erroneous self-reported dates were categorised as “Missing”.

### 2.7. Statistical Analysis

We summarised the sociodemographic and clinical characteristics at baseline using descriptive statistics, stratified by wave and migration status.

Analysis was conducted separately for each wave in the UK then meta-analysed. Participants were followed up from the start of each wave. Migrants who entered the UK after the start of a wave were followed up from the self-reported date of entry. If participants were not in the country (i.e. had not migrated to England) for the whole duration of a wave, they were not included in the analysis for that particular wave. Participants were censored either upon death or at the end of the study period by virtue of not meeting other censoring criteria. Participants were not censored at loss-to-follow-up due to the availability of linked data. The end of each period’s follow-up was the last date of hospitalisation during each wave (wave 1: June 10 2020, wave 2: March 28 2021, wave 3: October 25 2021).

We calculated the overall COVID-19-related hospitalisation incidence rates across the three waves and by wave and migration status. To estimate the total effect of migration status on the rate of COVID-19-related hospitalisation, we applied fixed-effect Poisson regression for wave 1 and fixed-effect negative binomial regression for waves 2 and 3. We found no evidence of multicollinearity. Multivariable adjustment was conducted using age at baseline, sex at birth, and minority ethnicity status. Models from the three waves were pooled using meta-analysis.

Baseline and weekly survey response data were extracted from REDCap, linked and analysed in R (version 4.1.2) and RStudio (version 1.4.1103).

### 2.8. Sensitivity analyses

We conducted two sensitivity analyses. In the main analysis, we carried out a complete case analysis. In the first sensitivity analysis, we included participants with missing country of birth under a “Missing” category. Ethnicity was considered an a priori confounder of the effect of migration status on COVID-19-related hospitalisation. However, the interplay between migration status and ethnicity may result in overadjustment for the effect of migration status on COVID-19-related hospitalisation when ethnicity is included as a confounder24. Thus, we carried out a sensitivity analysis without adjusting for ethnicity.

## 3. Results

Across the three waves, there were 148 counts of COVID-19-related hospitalisations. The overall estimated incidence rate of COVID-19-related hospitalisations was 3.0 per 1,000 person-years [95% CI 2.5-3.5]. When stratified by migration status, the incidence rates were 2.7 [2.2-3.2] and 4.6 [3.1-6.7] per 1,000 person-years in UK-born and migrant individuals, respectively.

Between March 1 and June 10 2020 (wave 1), 43 incident COVID-19-related hospitalisations were identified. The estimated incidence of COVID-19-related hospitalisation was 4.9 per 1,000 person-years [3.4-6.7] in UK-born participants and 6.7 per 1,000 person-years [2.7-13.8] in migrants.

Between September 1 2020 and March 28 2021 (wave 2), there were 76 counts of COVID-19-related hospital admissions. When stratified by migration status, the incidence rate was 4.1 per 1,000 person-years [3.1-5.2] in UK-born participants and 6.5 per 1,000 person-years [3.5-10.9] in migrants. Between June 1 and October 25 2021 (wave 3), there were 29 counts of COVID-19-related hospital admissions. Respectively, the incidence rate was 2.0 per 1,000 person-years [1.2-3.0] and 5.2 per 1,000 person-years [2.3-10.3] in UK-born individuals and migrants, respectively (Table 2).

**Table 1.** Demographic and clinical characteristics of the analysis cohort by wave. Follow up for wave 1 started from March 01 to June 10 2020, wave 2 from September 1 2020 to March 28 2021, and wave 3 from June 1 to October 25 2021.

| Characteristics | Wave 1 cohort<br>N=30,234 |  | Wave 2 cohort<br>N=30,275 |  | Wave 3 cohort<br>N=30,217 |  |
| --- | --- | --- | --- | --- | --- | --- |
|  | UK-born<br>N =26,492 | Migrant<br>N = 3,742 | UK-born<br>N =26,492 | Migrant<br>N =3,783 | UK-born<br>N =26,439 | Migrant<br>N=3,778 |
| **Age group** |  |  |  |  |  |  |
| 18-44 | 5,803<br>(22%) | 1,633<br>(44%) | 5,803<br>(22%) | 1,660<br>(44%) | 5,802<br>(22%) | 1,661<br>(44%) |
| 45-64 | 10,669<br>(40%) | 1,290<br>(34%) | 10,669<br>(40%) | 1,301<br>(34%) | 10,656<br>(40%) | 1,299<br>(34%) |
| 65+ | 10,020<br>(38%) | 819<br>(22%) | 10,020<br>(38%) | 822<br>(22%) | 9,981<br>(38%) | 818<br>(22%) |
| **Sex (at birth)** |  |  |  |  |  |  |
| Male | 11,277<br>(43%) | 1,543<br>(41%) | 11,277<br>(43%) | 1,562<br>(41%) | 11,246<br>(43%) | 1,561<br>(41%) |
| Female | 15,215<br>(57%) | 2,199<br>(59%) | 15,215<br>(57%) | 2,221<br>(59%) | 15,193<br>(57%) | 2,217<br>(59%) |
| **Household Region** |  |  |  |  |  |  |
| East Midlands | 2,480 | 145 | 2,480 | 145 | 2,479 | 145 |
|  | (9.4%) | (3.9%) | (9.4%) | (3.8%) | (9.4%) | (3.8%) |
| East of England | 6,107<br>(23%) | 543<br>(15%) | 6,107<br>(23%) | 549<br>(15%) | 6,094<br>(23%) | 548<br>(15%) |
| London | 2,861<br>(11%) | 1,831<br>(49%) | 2,861<br>(11%) | 1,857<br>(49%) | 2,857<br>(11%) | 1,853<br>(49%) |
| North East | 1,491<br>(5.6%) | 92<br>(2.5%) | 1,491<br>(5.6%) | 92<br>(2.4%) | 1,487<br>(5.6%) | 92<br>(2.4%) |
| North West | 3,205<br>(12%) | 147<br>(3.9%) | 3,205<br>(12%) | 147<br>(3.9%) | 3,195<br>(12%) | 147<br>(3.9%) |
| South East | 5,159<br>(19%) | 624<br>(17%) | 5,159<br>(19%) | 627<br>(17%) | 5,149<br>(19%) | 627<br>(17%) |
| South West | 2,050<br>(7.7%) | 158<br>(4.2%) | 2,050<br>(7.7%) | 161<br>(4.3%) | 2,047<br>(7.7%) | 161<br>(4.3%) |
| West Midlands | 1,517<br>(5.7%) | 90<br>(2.4%) | 1,517<br>(5.7%) | 91<br>(2.4%) | 1,513<br>(5.7%) | 91<br>(2.4%) |
| Yorkshire and<br>The Humber | 1,436<br>(5.4%) | 78<br>(2.1%) | 1,436<br>(5.4%) | 79<br>(2.1%) | 1,433<br>(5.4%) | 79<br>(2.1%) |
| Missing | 186<br>(0.7%) | 34<br>(0.9%) | 186<br>(0.7%) | 35<br>(0.9%) | 185<br>(0.7%) | 35<br>(0.9%) |
| **Years since migration upon start of their follow-up** |  |  |  |  |  |  |
| <1 | - | 122<br>(3.3%) | - | 101<br>(2.7%) | - | 43<br>(1.1%) |
| 1-5 | - | 420<br>(11%) | - | 434<br>(11%) | - | 396<br>(10%) |
| 5+ | - | 3,169<br>(85%) | - | 3,217<br>(85%) | - | 3,309<br>(88%) |
| Missing | - | 31<br>(0.8%) | - | 31<br>(0.8%) | - | 30<br>(0.8%) |
| **Ethnicity (binary)** |  |  |  |  |  |  |
| White British | 25,129<br>(95%) | 736<br>(20%) | 25,129<br>(95%) | 738<br>(20%) | 25,076<br>(95%) | 735<br>(19%) |
| Minority ethnic | 1,363<br>(5.1%) | 3,006<br>(80%) | 1,363<br>(5.1%) | 3,045<br>(80%) | 1,363<br>(5.2%) | 3,043<br>(81%) |
| **IMD Quintiles** |  |  |  |  |  |  |
| 1 (most<br>deprived) | 2,436<br>(9.2%) | 512<br>(14%) | 2,436<br>(9.2%) | 518<br>(14%) | 2,426<br>(9.2%) | 519<br>(14%) |
| 2 | 3,928<br>(15%) | 913<br>(24%) | 3,928<br>(15%) | 926<br>(24%) | 3,923<br>(15%) | 924<br>(24%) |
| 3 | 5,342<br>(20%) | 834<br>(22%) | 5,342<br>(20%) | 840<br>(22%) | 5,332<br>(20%) | 840<br>(22%) |
| 4 | 6,850<br>(26%) | 717<br>(19%) | 6,850<br>(26%) | 728<br>(19%) | 6,840<br>(26%) | 724<br>(19%) |
| 5 (least<br>deprived) | 7,750<br>(29%) | 732<br>(20%) | 7,750<br>(29%) | 736<br>(19%) | 7,733<br>(29%) | 736<br>(19%) |
| Missing | 186<br>(0.7%) | 34<br>(0.9%) | 186<br>(0.7%) | 35<br>(0.9%) | 185<br>(0.7%) | 35<br>(0.9%) |
| **Occupational risk** |  |  |  |  |  |  |
| No risk | 11,470<br>(43%) | 1,150<br>(31%) | 11,470<br>(43%) | 1,165<br>(31%) | 11,434<br>(43%) | 1,163<br>(31%) |
| Low risk | 4,212<br>(16%) | 854<br>(23%) | 4,212<br>(16%) | 861<br>(23%) | 4,209<br>(16%) | 860<br>(23%) |
| Elevated risk | 6,844<br>(26%) | 1,050<br>(28%) | 6,844<br>(26%) | 1,060<br>(28%) | 6,838<br>(26%) | 1,060<br>(28%) |
| High risk | 1,854<br>(7.0%) | 312<br>(8.3%) | 1,854<br>(7.0%) | 313<br>(8.3%) | 1,854<br>(7.0%) | 313<br>(8.3%) |
| Missing | 2,112<br>(8.0%) | 376<br>(10%) | 2,112<br>(8.0%) | 384<br>(10%) | 2,104<br>(8.0%) | 382<br>(10%) |
| **Clinical vulnerability** |  |  |  |  |  |  |
| Not clinically vulnerable | 14,577<br>(55%) | 2,270<br>(61%) | 14,577<br>(55%) | 2,301<br>(61%) | 14,564<br>(55%) | 2,300<br>(61%) |
| Clinically vulnerable | 8,314<br>(31%) | 992<br>(27%) | 8,314<br>(31%) | 996<br>(26%) | 8,283<br>(31%) | 994<br>(26%) |
| Clinically extremely vulnerable | 2,466<br>(9.3%) | 289<br>(7.7%) | 2,466<br>(9.3%) | 292<br>(7.7%) | 2,464<br>(9.3%) | 292<br>(7.7%) |
| Missing | 1,135<br>(4.3%) | 191<br>(5.1%) | 1,135<br>(4.3%) | 194<br>(5.1%) | 1,128<br>(4.3%) | 192<br>(5.1%) |
| **Living with 5-17 year old children** |  |  |  |  |  |  |
| Yes | 3,732<br>(14%) | 844<br>(23%) | 3,732<br>(14%) | 851<br>(22%) | 3,731<br>(14%) | 852<br>(23%) |
| No | 22,760<br>(86%) | 2,898<br>(77%) | 22,760<br>(86%) | 2,932<br>(78%) | 22,708<br>(86%) | 2,926<br>(77%) |
| **Total number of COVID-19 vaccination doses received** |  |  |  |  |  |  |
| 0 | 26,492<br>(100%) | 3,742<br>(100%) | 4,539<br>(17%) | 1,312<br>(35%) | 420<br>(1.6%) | 117<br>(3.1%) |
| 1 | - | - | 20,000<br>(75%) | 2,186<br>(58%) | 237<br>(0.9%) | 69<br>(1.8%) |
| 2 | - | - | 1,953<br>(7.4%) | 285<br>(7.5%) | 25,782<br>(98%) | 3,592<br>(95%) |

**Table 2.** Crude COVID-19-related hospitalisation rates by wave and by migration status. Number of COVID-19-related hospital admission and total person-years at risk omitted to avoid disclosure.

| Characteristics | N (%) | Mean follow-up time<br>(standard deviation) | Incidence rate (95%<br>CI) per 1,000 person-years |
| --- | --- | --- | --- |
| **Overall** |  |  |  |
| All | 30,276 | 604.1 (15.3) | 3.0 (2.5, 3.5) |
| UK-born | 26,492 (87.5%) | 604.4 (12.4) | 2.7 (2.2, 3.2) |
| Migrant | 3,784 (12.5%) | 601.8 (28.0) | 4.6 (3.1, 6.7) |
| **Wave 1** |  |  |  |
| All | 30,234 | 102.0 (0.8) | 5.1 (3.7, 6.9) |
| UK-born | 26,492 (87.6%) | 102.0 (0.0) | 4.9 (3.4, 6.7) |
| Migrant | 3,742 (12.4%) | 101.9 (2.4) | 6.7 (2.7, 13.8) |
| **Wave 2** |  |  |  |
| All | 30,275 | 208.8 (4.4) | 4.4 (3.5, 5.5) |
| UK-born | 26,492 (87.5%) | 208.9 (3.7) | 4.1 (3.1, 5.2) |
| Migrant | 3,783 (12.5%) | 208.4 (7.9) | 6.5 (3.5, 10.9) |
| **Wave 3** |  |  |  |
| All | 30,217 | 147.9 (3.1) | 2.4 (1.6, 3.4) |
| UK-born | 26,439 (87.5%) | 147.9 (3.2) | 2.0 (1.2, 3.0) |
| Migrant | 3,778 (12.5%) | 147.9 (2.0) | 5.2 (2.3, 10.3) |

Pooled unadjusted incidence rate ratio (IRR) suggested a 1.68 [1.08-2.60] increased rate of COVID-19-related hospitalisation in migrants compared to UK-born individuals (Figure 2). After adjusting for age, sex and ethnic minority status, COVID-19-related hospitalisation rate was higher in migrants than UK-born participants (pooled adjusted IRR: 1.35 [0.71-2.60]) (Figure 2), but reduced compared to unadjusted estimates. Country of birth was unknown for 2,805 individuals. In pre-specified sensitivity analyses, we found a consistent effect of migration status on COVID-19-related hospitalisation with analysis on the cohort including those with missing country of birth in the “Missing” category (pooled adjusted IRR: 1.55 [0.83-2.89]). When ethnicity was excluded as a confounder, the pooled adjusted IRR for migrants was 1.88 [1.20-2.95], suggesting an increased rate of COVID-19-related hospitalisation in migrants compared to UK-born individuals across the three waves (Supplementary Figure 2).

**Figure 1.**
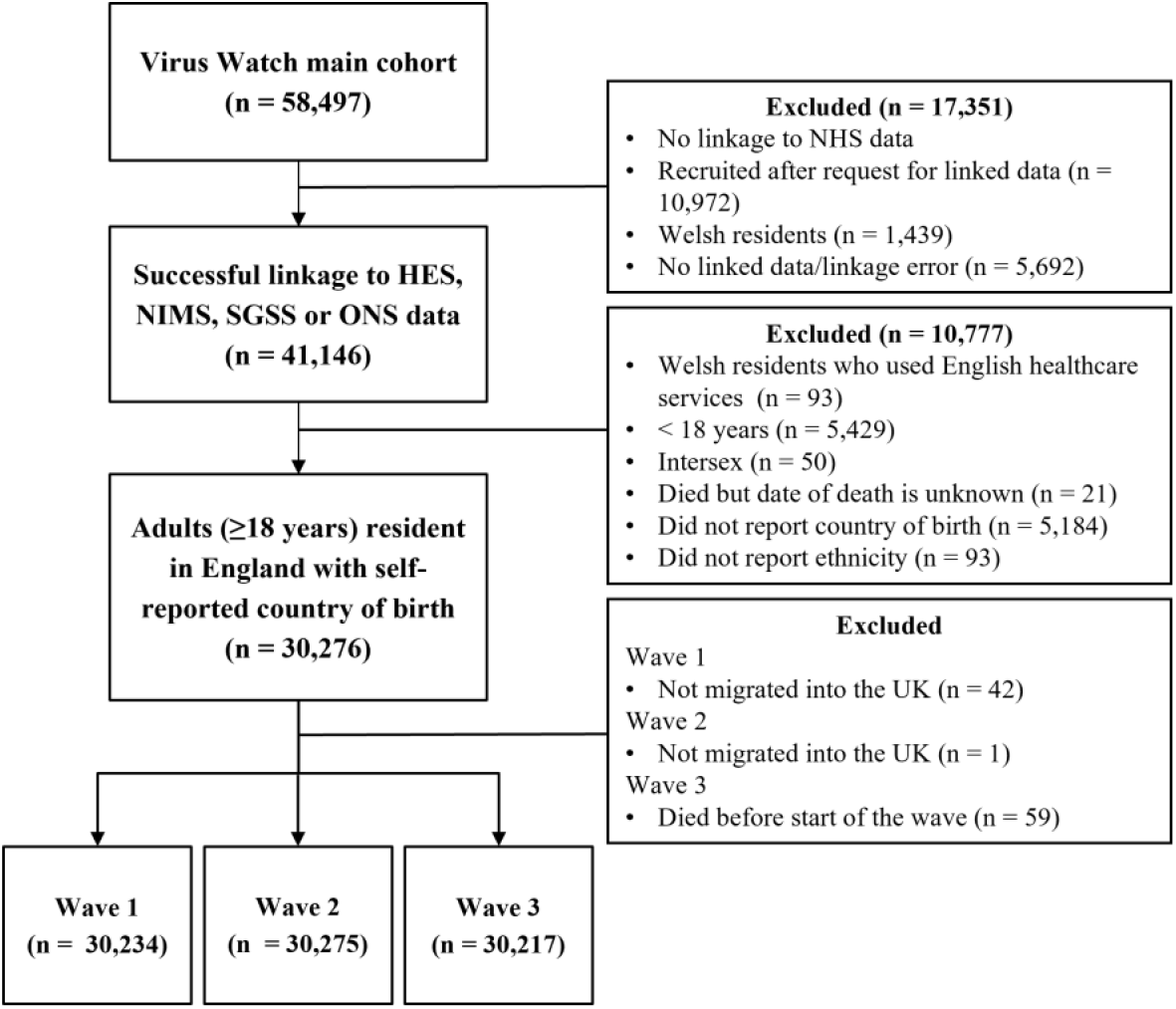
Flow diagram of participant eligibility. Selection of participants for inclusion in the present study is illustrated in Figure 1. As of March 2022, from a total of 58,497 Virus Watch participants, 41,146 (70.3%) were successfully linked to NIMS, HES, SGSS or ONS mortality data. 30,276 (73.6%) adults aged 18 years or older who met the selection criteria were included in this study. Table 1 reports the sociodemographic and clinical characteristics of the Virus Watch cohort participants included in this analysis by wave and migration status (wave 1: n=30,234; wave 2: n=30,275; wave 3: n=30,217). The combined cohort comprised 26,492 (87.5%) UK-born participants and 3,784 (12.5%) migrants. Over 80% of the migrants have been living in the UK for 5+ years upon the start of their follow-up. Compared to UK-born individuals, migrants were younger, more likely to be an ethnic minority, less clinically vulnerable, more likely to live in more deprived neighbourhoods and have high risk occupations (Table 1).

**Figure 2.**
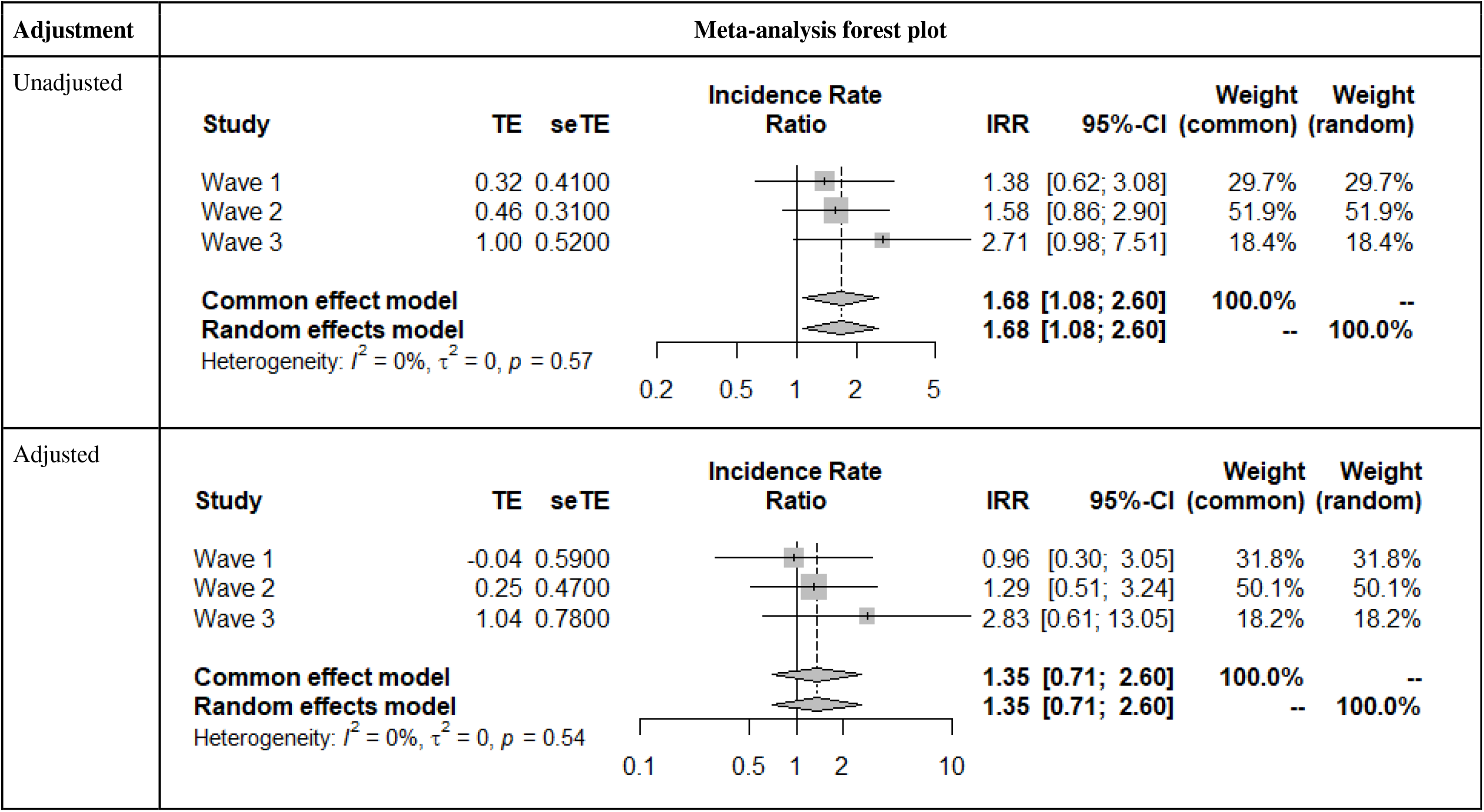
Association between migrant status and COVID-19-related hospitalisation in England – unadjusted and adjusted pooled incidence rate ratios (IRR). For all analyses, ‘UK Born’ was used as a reference category. Pooled IRR < 1 suggests decreased rate of COVID-19-related hospitalisation, pooled IRR = 1 suggests no difference in rate of COVID-19-related hospitalisation, pooled IRR > 1 suggests increased rate of COVID-19-related hospitalisation.

## 4. Discussion

This study estimated the incidence rates of COVID-19-related hospitalisation by wave and migration status and the total effect of migration status on the hospitalisation rate across COVID-19 infection waves 1-3 in England. Across the three waves, the crude COVID-19-related hospitalisation incidence rates were consistently higher in migrants than UK-born individuals in our primary and sensitivity analyses.

Migrants had higher hospitalisation rates when controlling for key variables but when ethnicity was adjusted for, these higher rates were reduced and became non-significant suggesting similar issues may account for higher rates in both ethnic minority and migrant groups. This corroborates a large-scale UK study that found greater risk of severe COVID-19 in some minority ethnic populations compared to the White population [25]. Higher COVID-19-related hospitalisation rates in both ethnic minority and migrant groups highlight the need to tackle the inequalities that these populations face.

While our study lacked statistical power to demonstrate a significant difference in the rate of COVID-19-related hospitalisation between migrants and UK-born individuals, the point estimate of the pooled adjusted IRR suggests that migrants may have an increased hospitalisation rate than those UK-born. This may be influenced by lower COVID-19 vaccine uptake in migrants compared to UK-born participants, which is consistent with the reporting of under-immunisation of routine and COVID-19 vaccines among migrants in the UK and other European countries (Table 1) [3], [18]–[20]. Therefore, our findings underscore the importance of interventions that support inclusive vaccination campaigns that target migrant communities [19], [26], [27]. A higher proportion of migrants in our analysis cohort having occupations with high SARS-CoV-2 infection risk (e.g. healthcare and service occupations) compared to UK-born participants may also contribute to migrants’ increased rate of COVID-19-related hospitalisation. Particularly during wave 3, following the relaxation of pandemic restrictions (i.e. no lockdowns), migrants with precarious and public-facing work were more likely to have an increased exposure to SARS-CoV-2 due to their inability to work from home. This is supported by a previous Virus Watch analysis demonstrating that healthcare and other public-facing sectors remained or became more exposed to SARS-CoV-2 after lockdowns were lifted compared to other occupations [7].

Migrants diagnosed with COVID-19 are exempt from healthcare charges [28], but may not be aware of these exemptions, which may result in delayed attempts to seek care when sick [29]. Others may fear the charge being imposed through a lack of diagnosis due to limited testing opportunities. This may indicate barriers to secondary care among migrants as a result of lower levels of language proficiency, barriers in accessing reliable information and appropriate services.

To our knowledge, this is the first study in the UK to examine the association between migration status and COVID-19-related hospitalisation in England across multiple waves of the pandemic. Using linked HES APC records that encompass data from the start of the pandemic reduced the risk of recall bias and enabled stratified analysis by waves of infection. Stratifying by waves allowed us to adjust for time-varying confounding such as hospital bed availability, emergence of new SARS-CoV-2 variants and implementation of public health measures (e.g. lockdowns and vaccine programmes) and to assess how the effect of migration status changed over the three waves.

Several limitations are important when interpreting our findings. The Virus Watch cohort is not representative of the migrant population in England or have similar socio-demographic characteristics as those in other studies. Virus Watch is limited by the fact that only households with a lead householder able to speak English and access the internet were able to take part. Our analysis lacked power to demonstrate a statistical difference in the rate of COVID-19-related hospitalisation between migrants and non-migrants in England, however, all estimates showed an increased risk for migrants. More marginalised and less well-established migrant groups, whose risk factors and barriers are more pronounced (likely resulting in an increased risk of COVID-19-related hospitalisation), were also underrepresented, potentially due to factors including their economic situation, fear of confrontation with legal authorities, and linguistic and cultural barriers [30]. Thus, Virus Watch migrant participants may be less deprived and less likely to have insecure immigration status than a fully representative sample of UK-based migrants. Our study was also at risk of survivor bias e.g. if individuals who survived severe COVID-19 infections and signed up to Virus Watch were disproportionately UK-born. The aforementioned biases are all likely to contribute to an underestimation of the effect of migration status on COVID-19-related hospitalisation rate.

Our findings suggest the need for better prevention of SARS-CoV-2 infection and early access to healthcare for migrants who are infected. Preventative efforts should focus on increasing vaccination uptake in order to protect migrants from COVID-19-related hospitalisation. Good practices seen during the pandemic include providing free access to the COVID-19 vaccine to everyone, regardless of immigration status, NHS number or identity document, and setting up innovative vaccination access points in convenient areas with free extended hours. In addition, policymakers should fund the collaboration between local councils and trusted community partners to engage and build trust with migrant communities. This will aid the dissemination of accurate public health information, and allow policymakers to better understand the existing barriers that these communities face in order to inform the designing of targeted public health services. Continued efforts should be made to reduce occupational risk of COVID-19 for migrants to prevent infection. To reduce the risk of hospitalisation amongst those who are infected, further efforts should ensure that barriers to accessing care are removed, including access to primary care, and culturally and linguistically appropriate services [26].

## 5. Conclusion

As we continue to live with SARS-CoV-2, it is essential to study the causal mechanisms which may underlie differences in COVID-19-related hospitalisations observed in migrants as compared to UK-born individuals. This will enable better targeted policy recommendations to protect those who face the highest risk of severe disease in future waves and pandemics.

## Data Availability

Data from the Virus Watch cohort are available on ONS Secure Research Service. The data are available under restricted access as they contain sensitive health data. Access can be obtained by ONS Secure Research Service.

## Additional Information

### Ethics and Consent

The Virus Watch study was approved by the Hampstead NHS Health Research Authority Ethics Committee: 20/HRA/2320, and conformed to the ethical standards set out in the Declaration of Helsinki. All participants provided informed consent for all aspects of the study. Approval was also obtained from the Independent Group Advising on the Release of Data (IGARD) to use the NHS Digital data under DSA DARS-NIC-372269-N8D7Z-v1.6

### Contributors

Conceptualisation: RWA, WLEF; Methodology: WLEF, VGN, SB, YB, RWA; Data curation: WLEF, VGN, CG, AMDN, SB, TEB; Formal analysis: WLEF, VGN; Writing – original draft preparation: WLEF; Writing – review and editing: WLEF, RB, YB, VGN, SB, TEB, IB, EF, CG, SH, JK, AMDN, YO, PP, ST, AY, ACH, RWA; Visualisations: WLEF; Project administration: JK, VGN, SB, TEB, AMDN, RWA, ACH; Funding acquisition: RWA, ACH.

## Acknowledgments

This work was supported by the Medical Research Council [Grant Ref: MC_PC 19070] awarded to UCL on 30 March 2020 and Medical Research Council [Grant Ref: MR/V028375/1] awarded on 17 August 2020. The study also received $15,000 of advertising credit from Facebook to support a pilot social media recruitment campaign on 18th August 2020.

This study was supported by the Wellcome Trust through a Wellcome Clinical Research Career Development Fellowship to RWA [206602]. IB is supported by an NIHR Academic Clinical Fellowship. SB and TB are supported by an MRC doctoral studentship (MR/N013867/1).

From 1 May 2022 Virus Watch received funding from the European Union (Project: 101046314). Views and opinions expressed are however those of the author(s) only and do not necessarily reflect those of the European Union or the European Health and Digital Executive Agency (HaDEA). Neither the European Union nor the granting authority can be held responsible for them.

