## Supplementary figures and images for "The incidence of COVID-19-related hospitalisation in migrants in the UK: Findings from the Virus Watch prospective community cohort study"

### Supplementary Figure 1

**Supplementary Figure 1. Directed acyclic graph**

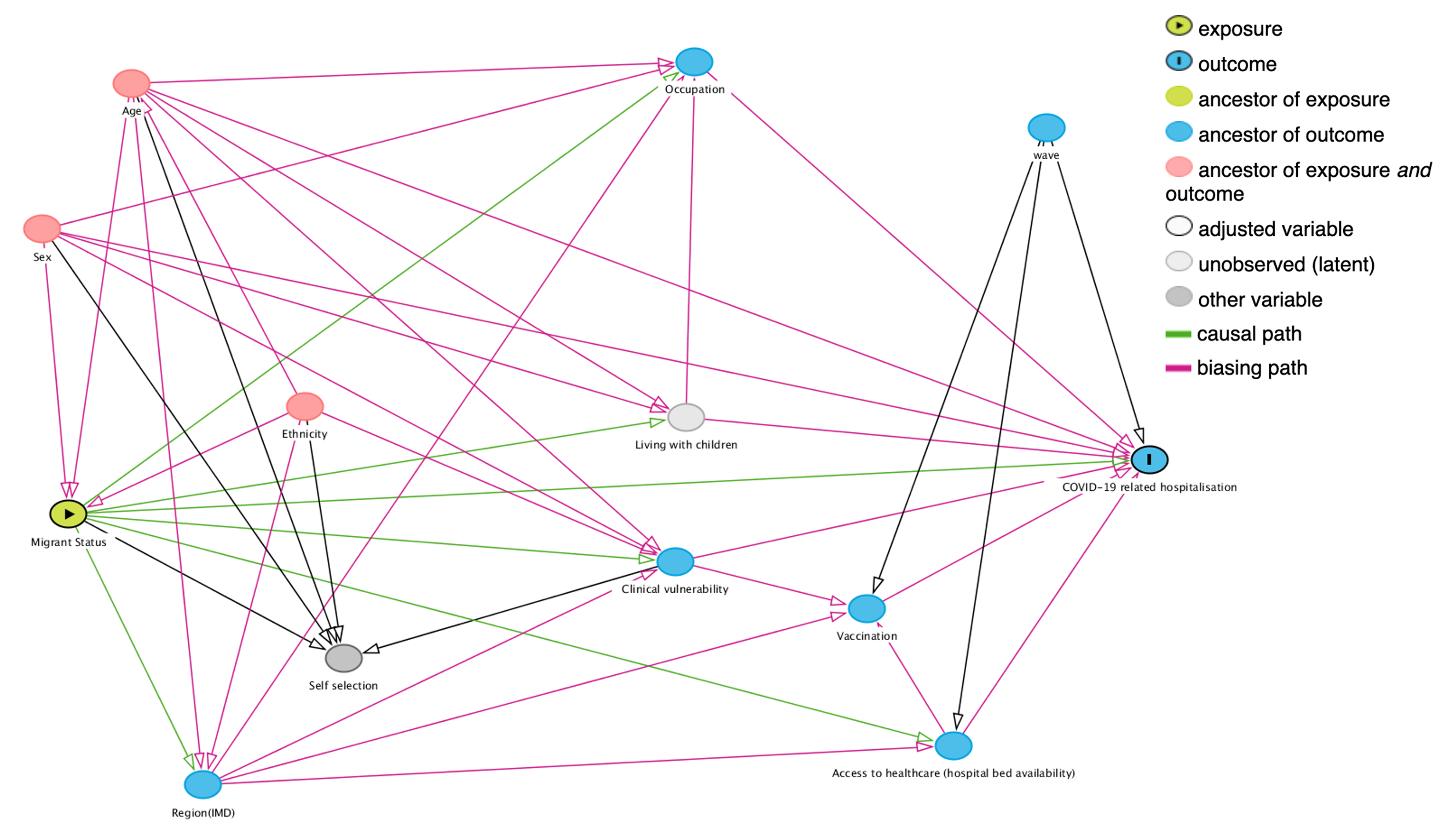
