## Supplementary Figure 2 for "The incidence of COVID-19-related hospitalisation in migrants in the UK: Findings from the Virus Watch prospective community cohort study"

**Supplementary Figure 2.  Sensitivity analyses forest plots .** For all analyses, ‘UK Born’ was used as a reference category. Pooled IRR < 1 suggests decreased rate of COVID-19-related hospitalisation, pooled IRR = 1 suggests no difference in rate of COVID-19-related hospitalisation, pooled IRR > 1 suggests increased rate of  COVID-19-related hospitalisation.

| **Sensitivity analyses** | **N** | **Meta-analysis forest plot** |
| --- | --- | --- |
| Including participants with missing migration status | 33,081 | Migrants in reference to UK-born individuals:  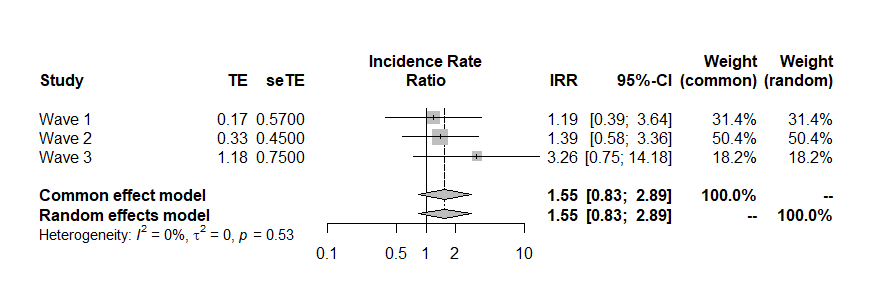  Individuals with missing migration status in reference to UK-born individuals: 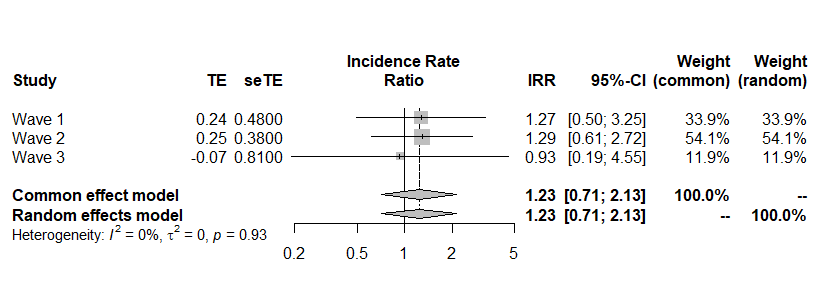 |
| Without adjusting for ethnicity | 30,369 | Migrants in reference to UK-born individuals:  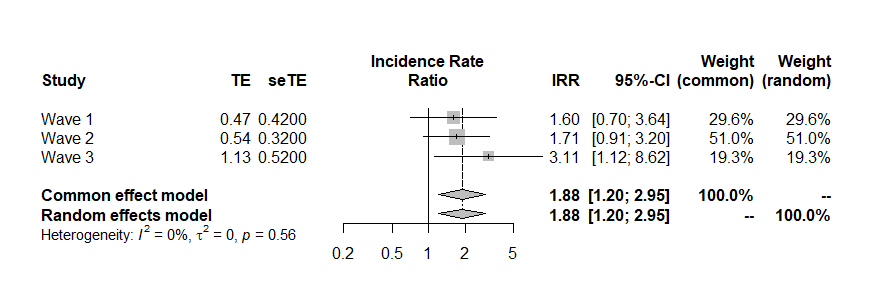 |
